# Increasing Cases of SARS-CoV-2 Omicron Reinfection Reveals Ineffective Post-COVID-19 Immunity in Denmark and Conveys the Need for Continued Next-Generation Sequencing

**DOI:** 10.1101/2022.09.13.22279912

**Authors:** Scott Burkholz, Michael Rubsamen, Luke Blankenberg, Richard T. Carback, Daria Mochly-Rosen, Paul E. Harris

## Abstract

SARS-CoV-2 has extensively mutated creating variants of concern (VOC) resulting in global infection surges. The Omicron VOC reinfects individuals exposed to earlier variants of SARS-CoV-2 at a higher frequency than previously seen for non-Omicron VOC. An analysis of the sub-lineages associated with an Omicron primary infection and Omicron reinfection reveals that the incidence of Omicron-Omicron reinfections is occurring over a shorter time interval than seen after a primary infection with a non-Omicron VOC. Our analysis suggests that a single infection from SARS-CoV-2 may not generate the protective immunity required to defend against reinfections from emerging Omicron lineages. This analysis was made possible by Next-generation sequencing (NGS), specifically of a Danish cohort with clinical metadata on both infections occurring in the same individual. We suggest that the continuation of COVID-19 NGS and inclusion of clinical metadata is necessary to ensure effective surveillance of SARS-CoV-2 genomics, assist in treatment and vaccine development, and guide public health recommendations.

## Main

The World Health Organization has designated five variants of concern (VOC) for SARS-CoV-2, the virus that causes COVID-19: Alpha, Beta, Gamma, Delta, and Omicron^1^. These VOC and emerging variants remain a significant obstacle in eliminating the SARS-CoV-2 pandemic^2^. The use of Next-generation sequencing (NGS) and accompanying metadata has allowed for a greater understanding of how VOC spread between hosts^3–5^. The Global Initiative on Sharing Avian Influenza Data (GISAID) has cataloged over 12 million SARS-CoV-2 sequences to date^3^. Data analysis suggests that each VOC is associated with a different level of infectivity and virulence^4,5^ and that novel variants, including currently circulating variants, have the potential to reinfect hosts despite global vaccination efforts^6,7^. The most recent variant, Omicron, or B.1.1.529, was designated in November 2021 as a VOC shortly after its initial identification in South Africa and Botswana^1^. The Omicron sub-lineages of BA.1, BA.2, and BA.3 have since spread around the world, becoming the initial dominant Omicron variants, followed by BA.4 and BA.5 in the weeks following^1,8^. With the exception of an Omicron-specific vaccine recently approved for use in the UK^9^ and US^10^, DNA, RNA, and whole protein vaccines designed utilizing original strain sequences have been used as part of the effort to control the pandemic. New cases of Omicron infections have exhibited levels of escape from vaccine induced neutralizing antibodies, leading to increased cases of reinfection^11,12^.

We report our analysis of the Danish COVID-19 Genome Consortium accessible through GISAID that examined individual cases of host reinfection across VOCs, including sub-lineages^3^. There are currently 21,708 reinfection entries available starting March 1st 2020 and ending August 28^th^ 2022. Each entry reports the exact collection date of both the initial infection and reinfection for the same individual, along with NGS sequences for the second infection. The primary infection and reinfection time-points were recorded as clinical metadata, allowing for the period between infections to be measured. A smaller portion of dataset entries (7595) reported the viral Pango lineage of both infections, in addition to the collection date, and NGS data for the initial infection and reinfection samples from the same individual. The Pango lineage nomenclature uses NGS to phylogenetically classify a virus based on its genomic composition^13,14^. The majority of the dataset did not have NGS data for the initial infection, and instead relied on a reverse transcriptase polymer chain reaction (rtPCR) assay to determine if the subject was positive for SARS-CoV-2 on the reported date. The rtPCR test does not specify the viral Pango lineage, and therefore those cases were not included in the analysis shown in Figure 1, but were included with NGS data in Figure 2a. 70 cases (<1% of total cases) had identical Pango lineages for the initial infection and reinfection. These cases were removed from the dataset because those entries would also be consistent with persistent and unresolved infection rather than reinfection.

**Figure 1:**
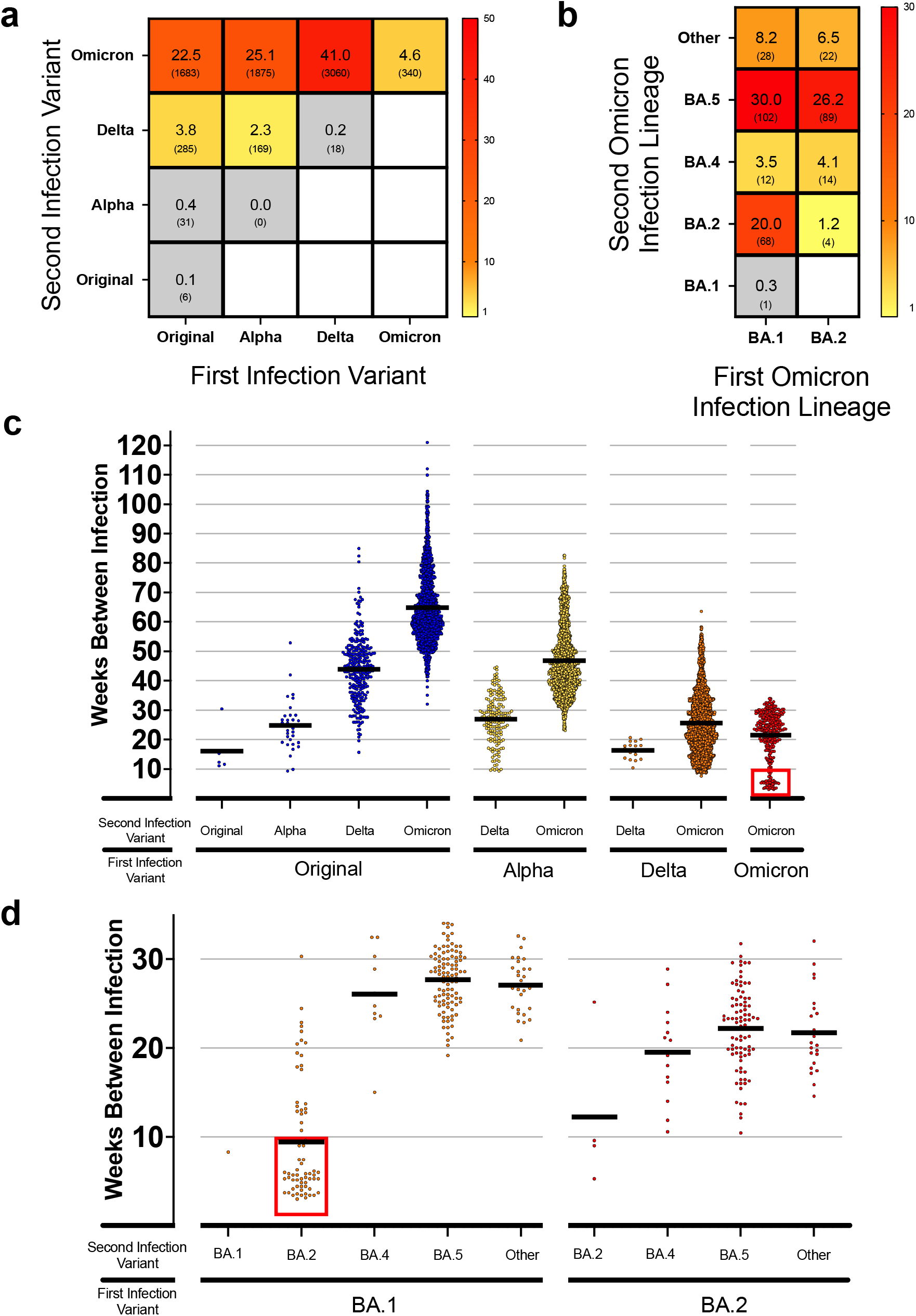
Reinfection Case Percent and Intervals by Variant. A) Heatmap showing the frequency of total reinfections between two variants for both an initial infection and reinfection in Denmark. Raw counts shown below frequency value in parenthesis. No data was available for blank white squares. B) Heatmap showing the reinfection frequency between an initial Omicron infection and a second Omicron infection by sub-linage in Denmark. Raw counts shown below frequency value in parenthesis. No data was available for blank white squares. C) Scatterplot showing the time between cases (weeks) for the first and second infection of different variants in Denmark. The red square highlights a number of early Omicron-to-Omicron cases mentioned in the text that occur before a 10-week period. D) Scatterplot for the time between cases (weeks) for Omicron-to-Omicron infections by lineage in Denmark. The red square highlights early Omicron-to Omicron cases mentioned in the text that occur before a 10-week period for specific sub-lineage.

**Figure 2:**
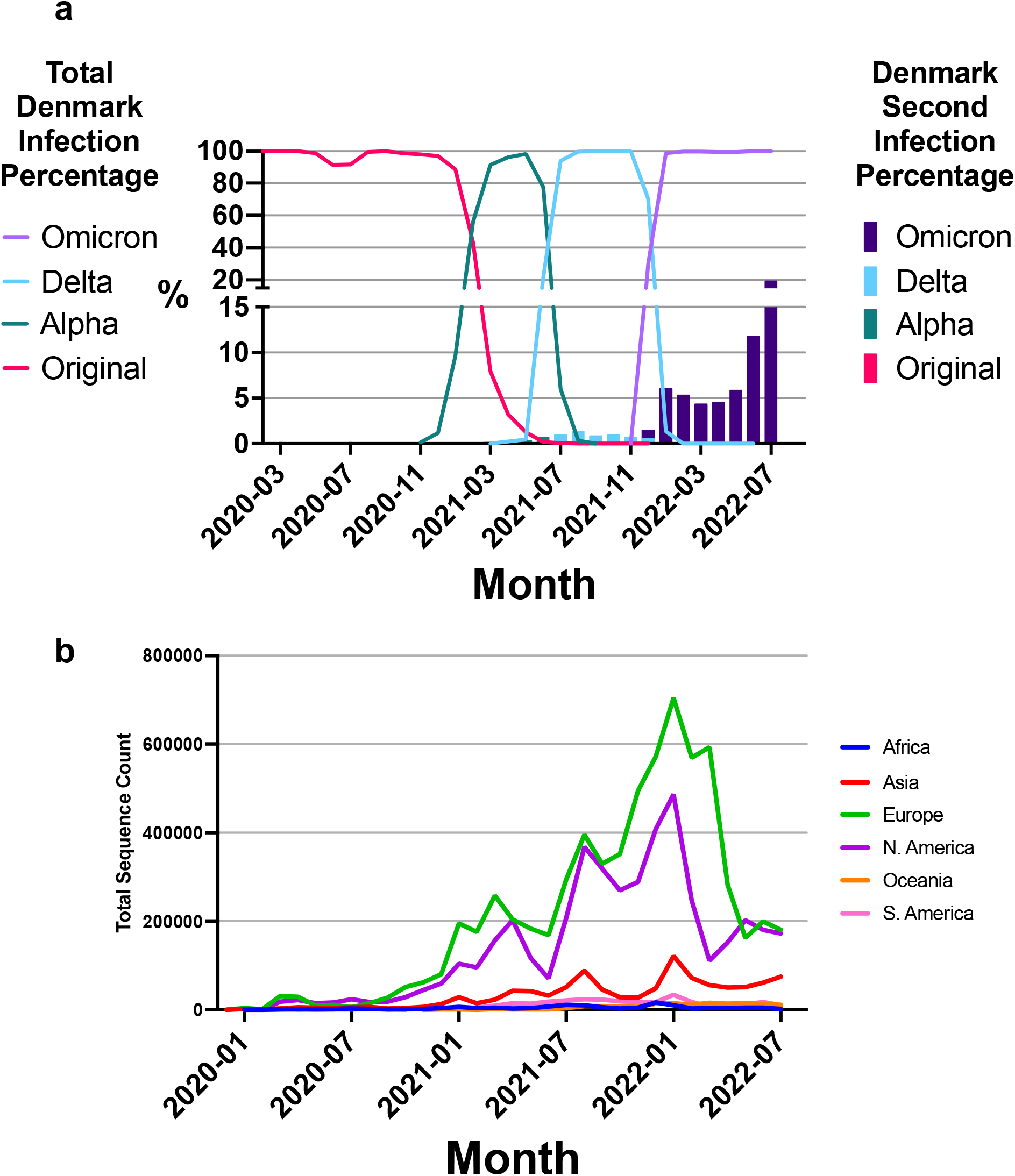
Reinfection Percentages and Sequencing Counts. A) Denmark sequencing percentages overtime stratified by variant represented as line plot with a bar plot overlay representing second infection percentages stratified by variant that include NGS and rtPCR data. B) Worldwide sequencing counts stratified by continent over a period of 1 month intervals.

For each VOC seen thus far, we found an increasing reinfection frequency favoring reinfection with the Omicron VOC (p<0.05, Chi-squared test) (Fig. 1a). 26% of individuals infected with the original viral strain showed increasingly higher reinfection frequencies with subsequent variants. Those initially infected with the Alpha variant had no cases of reinfection due to Alpha; however, increasing frequencies of reinfection from Delta (2.3%, 169 cases) and Omicron (25.1%, 1875 cases) were observed (Fig. 1a). For those initially infected with Delta, reinfection due to the Delta variant was limited (<1%, 18 cases), but 41% (3060 cases) were reported for Omicron variant reinfections. Thus far in the pandemic, reinfection within the same variant but different sub-lineages, other than for Omicron, was found to be small (0.3%, 24 cases), yet a higher number of individuals initially infected with Omicron report a reinfection due to Omicron sub-lineages (4.6%, 340 cases). There is a high frequency of reinfection with Omicron among all those reinfected since March 2020, during which time a total of 93.2% reinfections were due to Omicron. These results suggest that a primary infection with either the Original, Alpha, or Delta variant does not provide sufficient protection against reinfection, in particular for an Omicron reinfection.

In order to investigate this further, we stratified Omicron-to-Omicron reinfection cases by their major sub-lineage designation (Fig. 1b). The BA.2, BA.4, and BA.5 lineages closely share spike protein sequences compared to other Omicron lineages. There are only three mutational differences in the spike protein between BA.2 and both BA.4 and BA.5: del69-70, L452R, and F486V^8^. Despite this similarity, a high frequency of reinfections was reported (p<0.05, Chi-squared test) (Fig. 1b). Individuals initially infected with the BA.1 sub-lineage accounted for a high frequency of total Omicron-to-Omicron reinfections (62%, 211 cases) with the second infections predominately caused by BA.2 (20%, 68 cases) or BA.5 (30%, 102 cases) (Fig. 1b). Similarly, individuals initially infected with BA.2 showed comparably high frequencies of reinfection (38%, 129 cases) with BA.5 (26.2%, 89 cases) (Fig. 1b). Infections designated as “Other” in Figure 1b did not have a common nomenclature defining the lineage. The high frequencies of reinfection suggest that the characterized difference in the BA.1 or BA.2 and BA.5 spike protein is high enough to hinder the ability of post-infection neutralizing antibodies from BA.1 or BA.2 to bind to BA.5 spike protein^12,13^.

The decline of neutralizing antibodies following an infection raises a point of concern for how long natural immunity will last in a given individual. Pre-Omicron models estimated more than 90% effectiveness of initial post infection immunity, but these estimates decrease to less than 10% after 108 weeks^15^. Figure 1c and 1d show the time between infections by first and second VOC seen throughout the pandemic. Omicron-to-Omicron reinfections events appear in as little as 3 weeks after the initial infection, with a mean of 22 weeks (Fig. 1c). Of these Omicron-to-Omicron reinfections, 50 of the total 340 (14.7%) cases occur within 10 weeks of the initial infection; marked by a red box in Figure 1c. A t-test was conducted for the significance of the time between these Omicron-to-Omicron reinfections that occur before and after 10 weeks. These two sub-groups demonstrate significance of the mean reinfection times (p<0.0001, 95% CI: 17.17 - 20.19). Omicron-to-Omicron reinfections in Figure 1c are stratified by sub-lineage in Figure 1d. The reinfection occurrence before 10 weeks, marked by a red box in Figure 1d, is predominantly associated with BA.1 first and then reinfection with BA.2. Multiple t-tests (extended data table 1) between these groups in figure 1d shows significance between the difference in means for the majority of pairings.

Using a combined dataset of both NGS and rtPCR samples as described prior, a higher number of reinfections has been reported in the past eight months (December 2021 to July 2022) compared to other VOCs in the last year (Fig. 2a). During the global Omicron infection wave, Denmark had a 19.5% peak proportion of reinfection of the total sampled cases in July 2022. In comparison, a peak at 1.4% of total cases during the Delta wave were reported as reinfections. This difference highlights the high level of reinfections observed during the progression of the pandemic into the Omicron VOC.

Our results within this snapshot of available regional data illustrate two fundamental concepts. First, individuals are being reinfected with SARS-COV-2, and Omicron-to-Omicron reinfections appear to be occurring closer together with a higher frequency than seen for reinfections associated with previous VOC. Previous studies examine reinfection frequencies in SARS-CoV-2, including Omicron, but are often limited to PCR data only^7^, and do not designate the Pango lineage thus limiting the ability to gain insight into the pandemic. Second, there is an ongoing need for NGS with accompanying patient clinical metadata to continuously observe changes in viral infection rates and vaccine efficacy. Effective genomic tracking allows us to understand trends in viral infectivity and evaluate the current threat level of emerging VOCs.

We have seen an unprecedented level of worldwide scientific collaboration aimed at capturing viral sequences from infected individuals to build a public-facing database. This collaboration has made valuable data available to the public so that we can monitor changes in viral genomics, perform epidemiological studies, guide public health policy, and inform vaccine design^3^. Although GISAID has over 12 million sequences available at the time of this analysis, current sequencing rates are declining significantly^3^. A breakdown of NGS counts by continent is shown with recent declines in North America and Europe (Fig. 2b). This decline negatively affects the ability of our scientific community to analyze SARS-CoV-2 and COVID-19 as Pango lineage data can only be generated from NGS data. Furthermore, currently available genomic sequencing is often reported as either an initial infection or reinfection, but the initial infection and reinfection NGS data are typically not reported together for the same individual. The Denmark dataset illustrates the importance of reporting NGS data per individual, as a means to analyze reinfection risk. Another concern is breakthrough infections in vaccinated individuals from variants and their sub-lineages due to waning neutralizing antibody titers^16^. We could not explore this in our study specifically due to limited metadata. We strongly believe that Next-generation sequencing of SARS-CoV-2 samples must continue with addition of this metadata. In the case of reinfection, sequencing information characterizing both the timing between infections and viral genome composition of each infection is critical to inform accurate public health recommendations.

Our study suggests that the reinfections with the Omicron VOC are occurring at a higher frequency and over a shorter time interval than observed for other VOC earlier during the pandemic. This has significant implications for public health policy makers. The analysis presented here is an example of research that would not be possible without this robust dataset.

## Methods

### Data Retrieval

We utilized Next-generation sequencing metadata downloaded from the Global Initiate on Sharing Avian Influenza Data (GISAID, https://gisaid.org/) on August 28th, 2022^3^. The publicly available database is updated daily and was accessed at this time for two file sets. One GISAID file contains all current metadata for over 12 million sequences in the database and is cited in Supplementary Table 1. A second GISAID file set contains all filtered metadata of all people in Denmark that have two linked infections reported and is cited in Supplementary Table 2.

### Analysis

The metadata files are inputted into a custom written Python script to parse the large datasets and compute values of interest. The code is available at https://github.com/ and can be used to re-generate the data tables from the dataset. Entries with NGS were removed if Pango lineage for both infections is identical for the full, multi-digit designation. For these cases, it was difficult to discriminate between whether an individual experienced two distinct infections or a singular prolonged infection where they remained sick for an abnormal amount of time^17^. If the Pango lineage could not be resolved, it was also removed. After this filtering, Omicron sub-lineages are then converted to their major lineage such as BA.2.5 would become BA.2. Data on the VOC Gamma and Beta as well as “Other” variants without generally accepted nomenclature are removed from the dataset or not shown where applicable due to low levels of available sequences for analysis.

### Visualization

The data tables from the resulting code were input into GraphPad Prism (GraphPad Software Inc., San Diego, CA) for visualization through various plot types and formatting options.

### Statistics

Calculations for statistical significance was implemented by GraphPad Prism (GraphPad Software Inc., San Diego, CA) using a Chi-square test for independence and t-test for group mean comparison. Chi-square was performed on raw counts of cases for first and second infections as shown in Figures 1a and 1b. Raw case counts with a value of 0 were changed to 1 in order to perform this statistical test. Multiple t-tests were performed on select infection interval data as mentioned in the text shown in figure 1c and 1d. The t-test were performed under the conditions of unpaired, parametric, two-tailed, and a 95% confidence interval. The confidence interval utilizes the mean of the difference between the two groups of the t-test.

## Data Availability

All data are contained in the manuscript.
Our data and scripts (with exceptions for GISAID restrictions, but including instructions on how to download) will also be published on gitlab at https://gitlab.com/flowpharma.

https://gisaid.org/

## Data Availability

The SARS-CoV-2 metadata used in the analysis described are available through Global Initiate on Sharing Avian Influenza Data (GISAID, https://gisaid.org/)^3^. Specific sequence metadata utilized are cited in Supplementary Table 1 and 2.

## Code Availability

The code written for the purposes described in the methods to parse the dataset is available at https://github.com/.

## Acknowledgements

We appreciatively acknowledge the contribution of data from the Global Initiate on Sharing Avian Influenza Data (GISAID, https://gisaid.org/) through various authors and laboratories, in particular Povilas Matusevičius and The Danish COVID-19 Consortium. Specific sequences and metadata utilized are cited in Supplementary Table 1 and 2.

## Extended Data

Extended Data Table 1 – t-test of Omicron-to-Omicron reinfection groups in figure 1d.

## Supplementary Information

Supplementary Table 1 – GISAID Citation Table (All Infections)

Supplementary Table 2 – GISAID Citation Table (Denmark Reinfections Only)

